# Determinants of hospital outcomes for COVID-19 infections in a large Pennsylvania Health System

**DOI:** 10.1101/2021.09.08.21263311

**Authors:** Pamela A. Shaw, Jasper B. Yang, Danielle L. Mowery, Emily R. Schriver, Kevin B. Mahoney, Katharine J. Bar, Susan S. Ellenberg

## Abstract

There is growing evidence that racial and ethnic minorities bear a disproportionate burden from COVID-19. Temporal changes in the pandemic epidemiology and diversity in the clinical course require careful study to identify determinants of poor outcomes.

We analyzed 6255 individuals admitted with PCR-confirmed COVID-19 to one of 5 hospitals in the University of Pennsylvania Health System between March 2020 and March 2021, using electronic health records to assess risk factors and outcomes through 8 weeks post-admission. Discharge, readmission and mortality outcomes were analyzed in a multi-state model with multivariable Cox models for each transition.

Mortality varied markedly over time, with cumulative incidence (95% CI) 30 days post-admission of 19.1% (16.9, 21.3) in March-April 2020, 5.7% (4.2, 7.5) in July-October 2020 and 10.5% (9.1,12.0) in January-March 2021; 26% of deaths occurred after discharge. Average age (SD) at admission varied from 62.7 (17.6) to 54.8 (19.9) to 60.5 (18.1); mechanical ventilation use declined from 21.3% to 9-11%.

Compared to Caucasian, Black race was associated with more severe disease at admission, higher rates of co-morbidities and low-income resident zip code. Between-race risk differences in mortality risk diminished in multivariable models; while admitting hospital, increasing age, admission early in the pandemic, and severe disease and low blood pressure at admission were associated with increased mortality hazard. Hispanic ethnicity was associated with fewer baseline co-morbidities and lower mortality hazard (0.57, 95% CI: 0.37, .087).

Multi-state modeling allows for a unified framework to analyze multiple outcomes throughout the disease course. Morbidity and mortality for hospitalized COVID-19 patients varied over time but post-discharge mortality remained non-trivial. Black race was associated with more risk factors for morbidity and with treatment at hospitals with lower mortality. Multivariable models suggest there are not between-race differences in outcomes. Future work is needed to better understand the identified between-hospital differences in mortality.

## Introduction

The virus SARS-CoV-2, and its associated clinical disease, COVID-19, has had an immeasurable global impact. In the US, the virus rapidly spread throughout the country and has maintained a heavy clinical impact since. As the US epidemic has evolved, however, some of the early reported manifestations have appeared to shift, due in part to spread into different populations, increasing ability to recognize and treat COVID-19, as well as improved therapeutic approaches.

COVID-19 is well recognized to disproportionately affect systemically marginalized communities, including African Americans, Hispanics, and Native Americans [1–4]. Once hospitalized, however, the roles of race, ethnicity and other social determinants are less clear. Prospective therapeutic clinical trials [5, 6] and observational studies [7, 8] have focused on short-term outcomes such as 30-day mortality or discharge rates in hospitalized patients. Observational studies have yielded conflicting messages regarding the role of race and other participant characteristics on outcomes in hospitalized patients. Ogedegbe et al [9] reported better outcomes for Blacks compared to Whites within the New York University Langone Health system, including reduced risk of severe disease and lower mortality in hospitalized patients; in contrast, Mendy et al found increased risk for Blacks for these outcomes in the University of Cincinnati health system [10], and a recent meta-analysis and systematic review of 50 studies from across the United States and the United Kingdom found Black race was not associated with mortality or increased risk of ICU admission in hospitalized patients [2]. Determining causal relationships between race or other complex sociodemographic patient characteristics and outcomes is challenging with observational data. Evolving trends in the characteristics of infected and subsequently hospitalized individuals, including demographics, severity of illness amongst those infected, and changes in practice patterns further complicate disentangling the independent factors that contribute to patient outcomes.

The disease course of COVID-19 varies considerably across individuals, even among those severe enough to be hospitalized, with some hospitalized patients recovering after a brief or extended stay, others dying in hospital, while still others experience a worsening of disease and possible death or readmission after initial discharge. In the large body of work appearing in the medical literature to date that examines COVID-19 outcome, multivariate analyses of patient outcomes have either focused on a single clinical outcome, such as 30-day mortality, or investigators have applied separate models for different aspects of the disease. This approach frequently requires ignoring information on several other observed clinical outcomes for individual patients, which may result in an analysis of an incomplete or overly simplistic summary of the disease course reliant on single or composite outcomes. Multistate models allow for incorporation of multiple outcomes per patient, with flexible multivariable modeling of the risk factors for each outcome, and are relatively straightforward in standard statistical software. With this approach we are able to conduct a more comprehensive assessment of how patient risk factors affect different aspects of the potentially complex disease course experienced by patients hospitalized with COVID-19.

We describe the experience of patients hospitalized for COVID-19 in the University of Pennsylvania Health System (UPHS) from March 2020 through March 2021. Using the electronic health record data from five hospitals, we sought to identify key determinants of clinical outcomes in the eight-week period following first hospitalization for COVID-19. Of particular interest were temporal trends in the severity of illness and mortality, post-discharge outcomes, and the role of race on clinical outcomes. These data demonstrate substantial evolution of outcomes over time and identify several important determinants of disease outcomes.

## Methods

### Data collection and study cohort

Retrospective electronic health records (EHR) data were obtained from hospitalized subjects with COVID-19 infection admitted prior to March 31, 2021 at one of five hospitals within UPHS, with follow-up through May 26, 2021. Three hospitals were within city limits of Philadelphia, of which two were tertiary referral centers and all had major training programs; two were in outlying non-urban areas. Characteristics of these hospitals are presented in S1 Appendix. We abstracted demographic information, vital sign measurements, body mass index (BMI), recent comorbidity diagnoses, and hospitalization outcomes. Socioeconomic status was assigned according to the median household income for the patient’s residential zip code from the U.S. Census Bureau’s American Community Survey (ACS) for 2014-2018 [11]. Prior to study start, this study was approved and deemed exempt by the University of Pennsylvania Institutional Review Board (UPenn IRB). A limited dataset was generated from the EHR using the minimum necessary protected health information that was authorized by the UPenn IRB.

Baseline comorbidities designated by the CDC as underlying conditions that may increase risk of severe illness from COVID-19 [12] were abstracted from the EHR for the year prior to admission and classified using the *International Classification of Diseases, 10^th^ Revision* (ICD-10). We considered diabetes (Type 1 or Type 2), obesity, chronic kidney disease, chronic liver disease, chronic respiratory disease, cardiovascular disease, cancer, and immune deficiency.

More specific categorizations were considered for cardiovascular disease (coronary artery disease, hypertension, congestive heart failure, and other) and chronic respiratory disease (COPD, asthma, and chronic oxygen requirement). Further details are in the S1 Appendix.

### Statistical analysis

Descriptive statistics were calculated for baseline characteristics at time of first COVID-19 hospital admission. The median value of vital signs on the closest day to admission with data was selected as the baseline value. Blood pressure was classified as low if systolic blood pressure (SBP) < 90 millimeters of mercury (mm Hg) or diastolic blood pressure (DBP) < 60 mm Hg; otherwise, subjects were classified as having high blood pressure if SBP > 130 mm Hg or DBP > 80 mm Hg. Associations between patient baseline characteristics were evaluated using the chi-squared test.

Patient outcomes included hospital discharge, readmission, and mortality within 56 days of first hospital admission. Cumulative incidence curves for mortality and discharge for the first hospital stay were computed [13]. Discharge and death were analyzed as competing risks, as death following COVID recovery is presumed to be occurring at a different rate than for patients requiring hospitalization. For this analysis, the discharge outcome of interest was defined as discharge followed by survival to day 56; patients who remained in hospital for longer than 56 days were censored at day 56.

A multi-state model was created to investigate the determinants of risk for each of the potential outcome (states) patients experience throughout the disease course and incorporated data from all events on a patient within the follow-up period. Separate multivariable Cox models were fit for each transition between the three states of hospitalization, discharge, and death, within 56 days of their first COVID-19 hospital admission. Transitions from hospitalization to discharge, hospitalization to death, discharge to hospitalization, and discharge to death were considered, with death as the only absorbing state.

Using this multi-state framework, two sets of predictors were used to create transition-specific Cox regression models using the ‘mstate’ (Version 0.3.1) and ‘survival’ packages in the R software [14–16]. The primary model (model 1) included baseline age, sex, race, ethnicity, income, BMI, blood pressure, admitting hospital, admission epoch (Mar-Apr 2020, May-June 2020, July-Oct 2020, Nov-Dec 2020, Jan-Mar 2021), and an indicator of admission to an intensive care unit at the first hospital admission in order to control for baseline COVID-19 severity. For the transitions beginning in the hospital, the multi-state approach also allowed for the inclusion of a variable indicating whether a specific hospital stay was a first or re-admission. ICU admission was defined as admission to the ICU or placement on mechanical ventilation on the day of admission, to account for the mixed function COVID-19 units established outside of traditional intensive care units. Because disease severity at admission could mediate the effect of comorbidities on patient outcomes, a second model (model 2) was fit with these same patient characteristics plus EHR-documented comorbidities instead of the ICU-level admission variable.

Statistical tests were two-sided and done at the 0.05 significance level. A complete case analysis approach was taken, as only 22/6255 (0.35%) of patients had missing values for any variable of interest. Analysis was done using R software (Version 4.0.2) [16].

## Results

Our study cohort included 6255 hospitalized patients with a total of 6953 admissions. Table 1 shows patient characteristics at admission. The overall median (IQR) age was 62.0 years (47.0, 75.0) and 48.1% were male. About half of the patients were Black (44.1%) and 46.0 % were White; 9.4% were of Hispanic/Latinx ethnicity. Among all hospitalized, 14.8% received ICU-level care on day of first admission. A majority of patients had resident zip codes with median income less than $75,000. Reflecting the local epidemic, admissions for COVID-19 surged in early spring of 2020, waned over the summer until September 2020 before increasing to a peak in December 2020, with subsequent declines continuing through March of 2021 (S1 Fig).

**Table 1.**
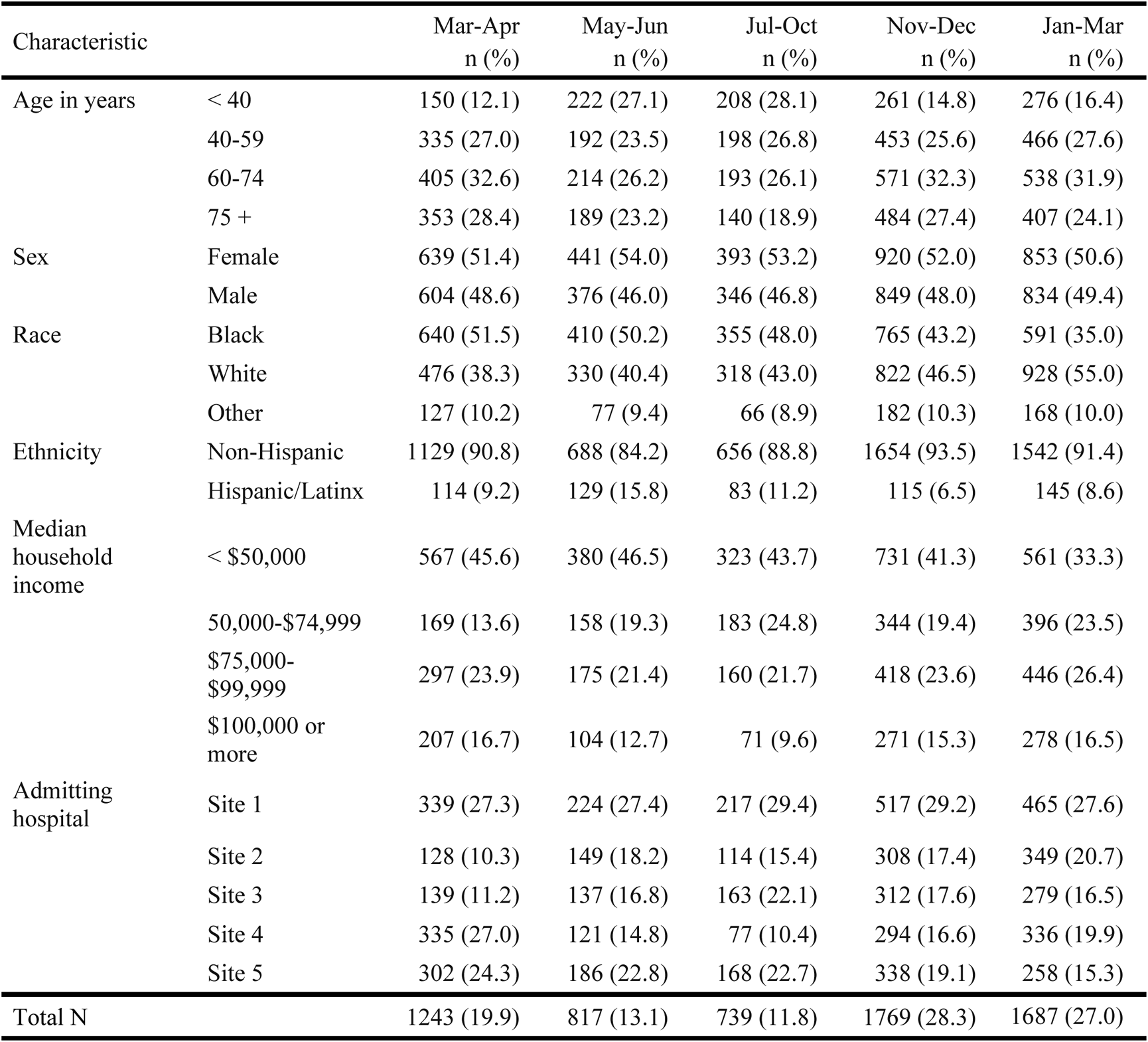
Patient characteristics at hospital admission for COVID-19 infection. N=6255. Overall chi-sq test p-values: p<0.001 for Age, Race, Ethnicity, Income, Entity; p=0.52 for Sex. Percentages may not add up to 100% due to missing data in income (n=15). Income: Median Household Income in patient’s 5-digit zip code, as determined by the 2014-2018 5-year ACS.

Table 1 shows time trends in patient demographics for COVID-19 hospital admissions. There was a marked shift in age, as younger patients (< 40 years) shifted from being the least common group in March-April 2020 to the most common group in July-October 2020, then returned to being the least common group in January-March 2021. There was also a notable shift in the racial distribution of hospitalized patients, as 51.5% of admitted patients in March-April 2020 identified as Black compared to only 35.0% in January-March 2021. Table 2 shows trends in severity of illness at presentation and reported comorbidities. In the later months, fewer individuals received ICU-level care on day of admission. The use of mechanical ventilation also decreased markedly; 9.4% of hospitalized patients in March-April 2020 received mechanical ventilation at admission, compared to 3.4% of patients admitted during January-March 2021, and the rates of mechanical ventilation at any time during the hospitalization declined from 21.7% to 10.7%. These time trends were similar across the five hospitals.

**Table 2.**
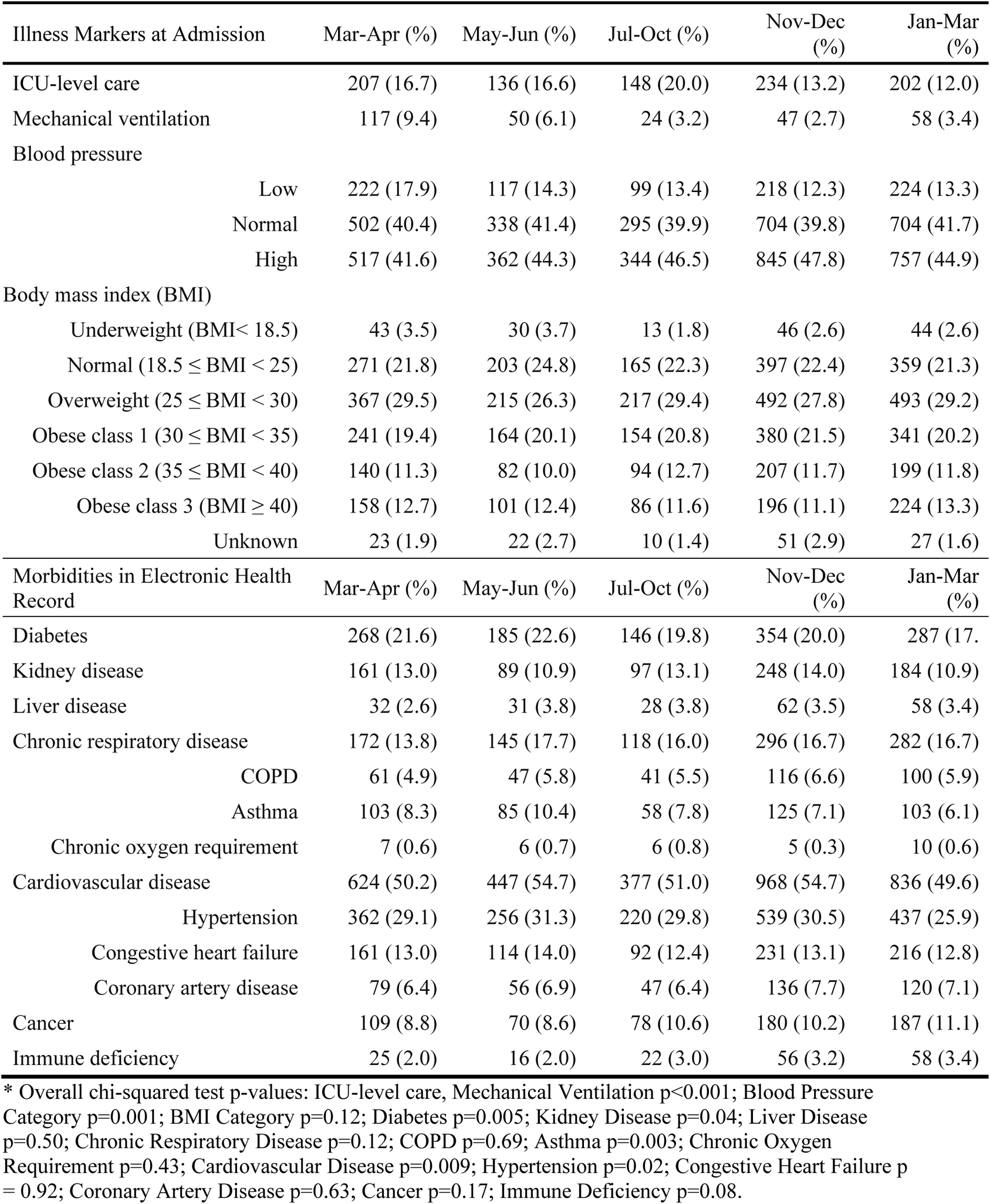
Severity of illness markers and comorbid conditions based on ICD-10 Codes recorded within one year prior to the COVID-19 hospital admission. N=6255.*

Many patients had relevant comorbidities at admission (Table 2). There were 72.8% who were obese or overweight, and 2.8% were underweight [17]. Over half of all patients (52.0%) had a history of cardiovascular disease. Chronic respiratory disease was present in 16.2% of patients. Diabetes (19.8%), kidney disease (12.5%), and cancer (10.0%) were also prevalent.

Comorbidities differed by race and ethnicity (S1 and S2 Tables). Black patients were significantly more likely to have at least one comorbid condition (71.3% versus 56.6% in Whites and 48.2% other races, p < 0.001). Diabetes, kidney disease, respiratory disease, and cardiovascular disease were significantly more common in Blacks than Whites and other races. Blacks were also more likely to require ICU-level care on admission (15.2% versus 13.6% for Whites and 18.8% for other races (p=0.002)). In contrast, Hispanic/Latinx patients were younger (38.3% versus 15.7% < 40 years) and less likely to have one or more comorbid conditions compared to non-Hispanic/Latinx (42.8% vs 64.2%, p<0.001).

Race was associated with residential zip code income (p < 0.001); 72.4% of Black patients resided in areas with median income < $50,000 compared to 13.7% for Whites and 27.5% for other races. Hispanic/Latinx patients resided in higher income areas compared to other ethnicities.

### Hospital outcomes

At eight weeks following initial COVID-19 admission, 797 patients (12.7%) had died, 5362 (85.7%) had been discharged without a recorded death, and 96 (1.5%) remained in hospital. Of the 797 deaths, 207 (26.0%) occurred after an initial discharge. Fig 1 shows the temporal trends in cumulative incidence curves for death and discharge, with overall incidence curves shown in S2 Fig. Mortality rates decreased significantly from March-April 2020 to July-October 2020 before rising slightly again in November 2020-March 2021. Conversely, hospital discharge rates increased significantly from March-October 2020 before dropping during the winter.

**Fig 1.**
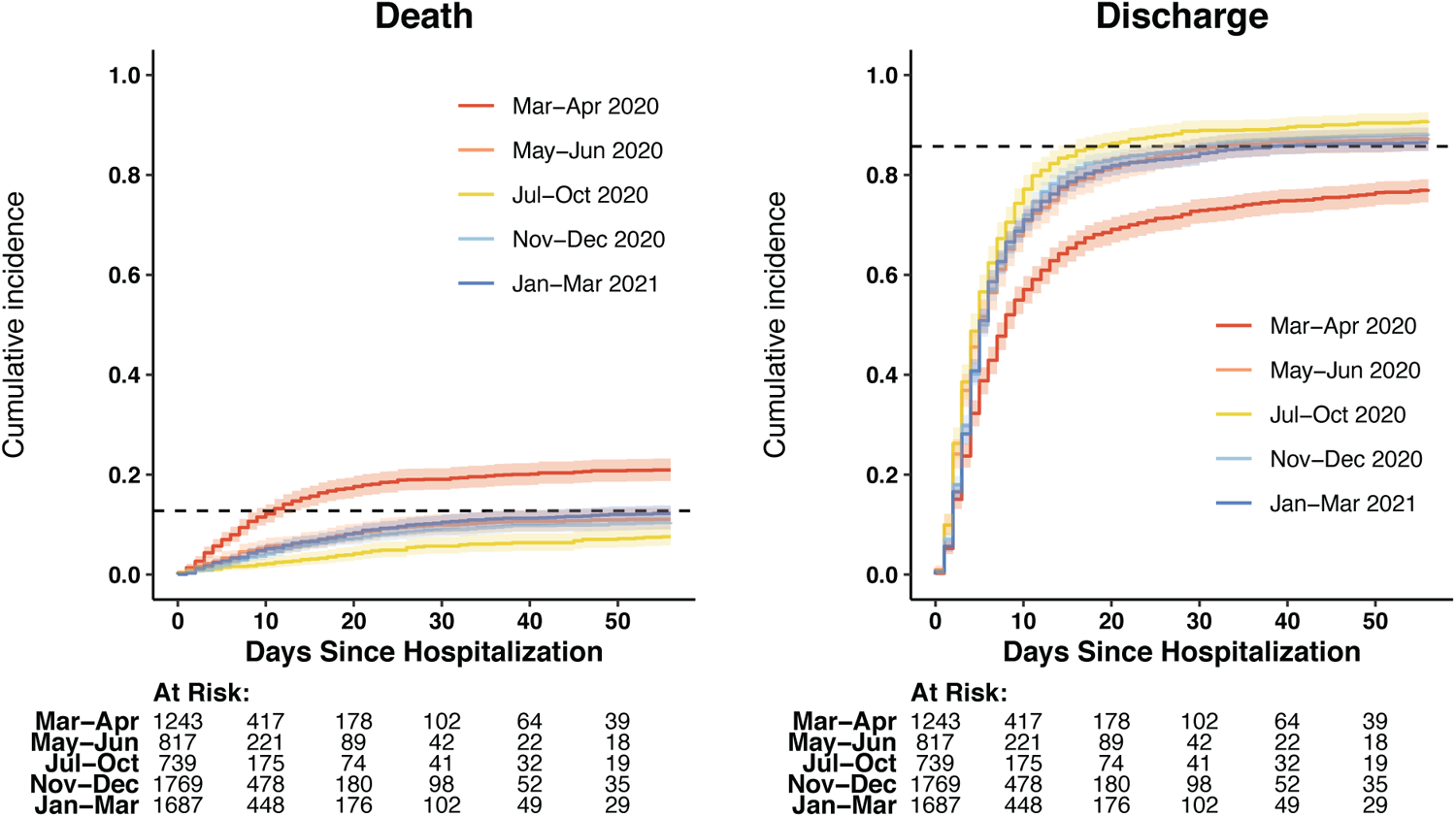
Cumulative Incidence Curves for Death and Discharge by Month. N = 6255.

Multivariable associations from the multistate models are discussed in detail below. Figs 2 and 3 and S3 and S4 Figs present the cause-specific hazard ratios (HRs) for our base model (model 1) that adjusted for baseline disease severity and S5–S8 Figs present the HRs for model 2 that adjusted for baseline comorbidities.

**Fig 2.**
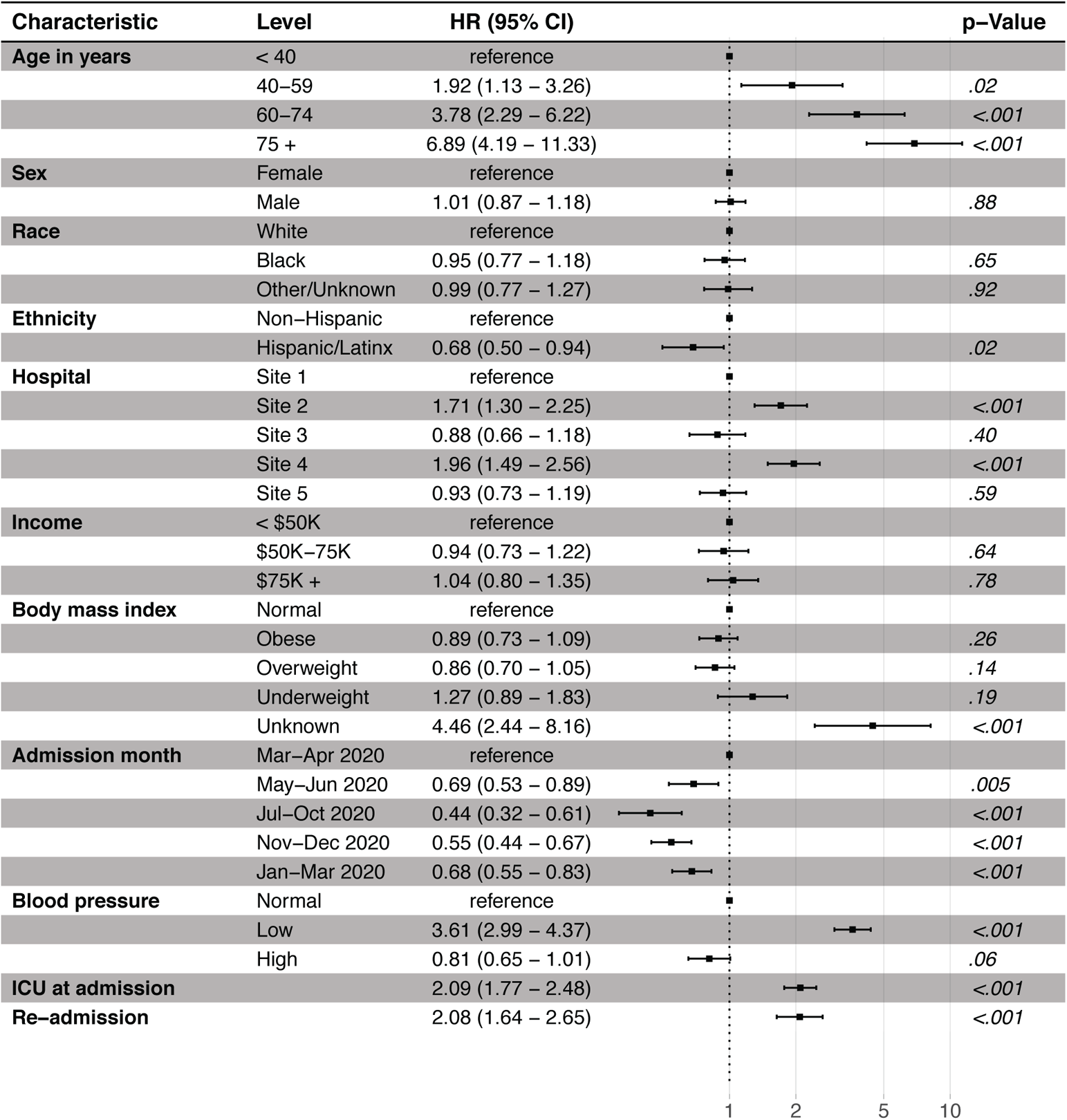
Hospitalization to death Cox regression (model 1). N=6233; 22 observations deleted due to missingness in zip code (15) and blood pressure (7). Site 1 (n=1762), Site 2 (n=1048), Site 3 (n=1030), Site 4 (n=1163), Site 5 (n=1252) are unique hospitals in the University of Pennsylvania Health System. Income: Median Household Income in patient’s 5-digit zip code, as determined by the 2014-2018 5-year ACS; CI: Confidence interval.

**Fig 3.**
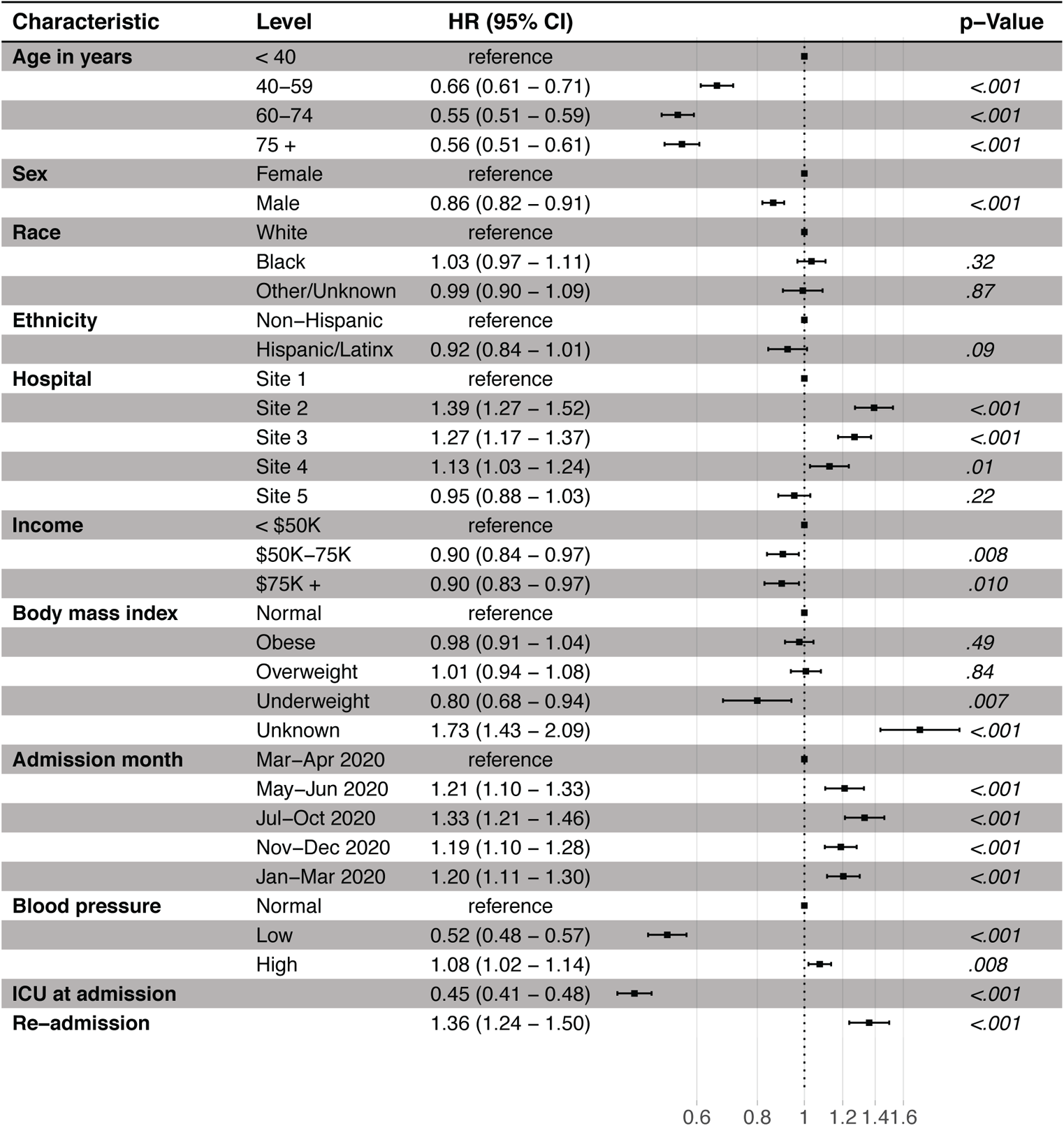
Hospitalization to discharge Cox regression (model 1). N=6233; 22 observations deleted due to missingness in zip code (15) and blood pressure (7). Site 1 (n=1762), Site 2 (n=1048), Site 3 (n=1030), Site 4 (n=1163), Site 5 (n=1252) are unique hospitals in the University of Pennsylvania Health System. Income: Median Household Income in patient’s 5-digit zip code, as determined by the 2014-2018 5-year ACS; CI: Confidence interval.

### Associations with in-hospital mortality

Fig 2 shows the HR for the hospitalization to death transition according to our base model (model 1), which adjusted for disease severity at admission (requiring ICU care) and other baseline characteristics. Death was strongly associated with age (p<0.001), with the HR steadily increasing with advanced age. Severe disease, captured by low blood pressure (HR 3.61, 2.99-4.37) and ICU-level care at admission (2.09, 1.77-2.48), and hospital re-admission (2.08, 1.64-2.65) were both associated with increased mortality hazard. Race, sex, and income were not predictors of mortality, while Hispanic/Latinx ethnicity was significantly associated with a lower HR (0.68, 0.50-0.94). The mortality hazard varied by admitting hospital, with admission to Site 2 (HR 1.71, 1.30-2.25) and Site 4 (1.96, 1.49-2.56), the two non-urban centers in our study, associated with increased mortality hazard compared to Site 1.

In our second model, which adjusted for comorbidities instead of ICU care at admission (model 2, S5 Fig), we did not find any additional associations with death. Other associations were similar to model 1.

### Associations with hospital discharge

Fig 3 presents the cause-specific hazard ratios for the hospitalization to discharge transition for model 1 and S6 Fig presents model 2. (Note that for hospital discharge, a higher hazard ratio reflects a relative benefit rather than a relative harm.) Age was again an important risk factor in both models (p <0.001); the discharge hazard decreased with older age, with the 75+ age group having a HR of 0.56 (95% CI: 0.51, 0.61) in model 1. In both models, male sex, underweight and low blood pressure were each associated with a decreased discharge hazard. Admission after March-April 2020 and unknown BMI were associated with an increased discharge hazard in both models. ICU at admission (0.45, 0.41-0.48) in model 1, and diabetes (0.93, 0.87-1.00), kidney disease (0.83, 0.76-0.90), chronic respiratory disease (0.91, 0.85-0.98), heart failure (0.70, 0.64-0.75), and cancer (0.85, 0.78-0.92) in model 2, were each associated with a decreased discharge hazard. History of hypertension (1.15, 1.08-1.23) and coronary artery disease (1.12, 1.01-1.25) were associated with and increased discharge hazard. Admitting hospital was also an important risk factor with Site 2, Site 3, and Site 4 all associated with an increased hazard for discharge in both models.

### Associations with death after discharge

Most patients (5569 of 6255, 89.0%) were discharged from their initial hospital admission within 56 days. Of the 797 deaths within the first 56 days of admission, 207 (19%) occurred after a first discharge. 102 (49.3%) of these 207 post-discharge deaths occurred during a readmission, while the other 105 occurred outside of a Penn hospital. Notably, 11.2% of the oldest patients (75 years-old and older) who were discharged died by day 56. In contrast, the rates of post-discharge deaths by Day 56 for ages 60-74, 40-59, and < 40 years were 2.7%, 1.0%, and 0.4%, respectively (p < 0.001). S3 Fig shows the cause-specific hazard ratios (HR) for the discharged to death transition according to our base model (model 1). Older age (HR 12.43, 1.64-94.3 for Age 60-74; 57.8, 7.87-424 for age 75+, each compared to age <40), underweight BMI (2.08, 1.08-4.02) and ICU during first admission (2.29, 1.48-3.55) were associated with an increased hazard for death, while obese BMI (0.39, 0.22-0.68) was associated with a decreased death hazard. The results from model 2 (S7 Fig) also revealed chronic respiratory disease (1.61, 1.01-2.55), heart failure (2.03, 1.26-3.26), and cancer (2.67, 1.68-4.24) as associated with an increased hazard for death.

### Hospital re-admissions

633 of the 5569 (11.4%) discharged patients were re-admitted at least once within 56 days of first admission, with some patients re-admitted three or more times. The median (IQR) time between admissions was 5 (1-12) days. S4 Fig shows the cause-specific hazard ratios for the discharged to hospitalization transition according to model 1. Older age (HR 1.48, 1.15-1.91 for ages 60-74; 2.43, 1.89-3.14 for 75+, each compared to age <40), male sex (1.17, 1.01-1.37), low blood pressure at first admission (1.41, 1.10-1.82), and ICU during first admission (1.83, 1.52-2.20) were each associated with an increased hazard for re-admission. Other/unknown race (0.53, 0.38-0.74 compared to White), admission to Sites 2, 3, 4, and 5 compared to Site 1, obese or overweight BMI, and admission after March-April 2020 were all associated with a decreased risk for hospital re-admission. S8 Fig shows the results for model 2 which, indicate that kidney disease (1.50, 1.22-1.85), chronic respiratory disease (1.67, 1.39-1.99), heart failure (1.68, 1.37-2.05), and cancer (1.51, 1.22-1.86) are all associated with an increased re-admission hazard.

### Outcomes by race and admitting hospital

Fig 4 (S9 Fig) shows the death (discharge) outcomes by race and admitting hospital. In all sites combined, the cumulative incidence curves suggest that Black race is associated with a decreased risk of death (Fig 4) and an increased probability of discharge (S9 Fig), but there are no apparent systematic differences by race when breaking down results by hospital. Black patients were admitted at a much a higher proportion to the urban Philadelphia sites (sites 1, 3, 5), which were associated with an overall lower hazard of death relative to the other two sites (Fig 2). Because the urban sites included the majority of patients and had a higher proportion of Black patients, the unadjusted comparison of outcomes by race shows overall better outcomes for Blacks compared to Whites (Fig 4). In our base multivariable model, there was no association between Black race and outcome for any of the transitions after adjustment for admitting hospital (Figs 2 and 3, S3 and S4 Figs). Adjusting for comorbidities in model 2 yielded similar results, although a small beneficial effect for black race remained marginally significant for the hospital to discharge and discharge to re-hospitalization transitions after adjusting for admitting hospital (S5–S8 Figs). In view of the multiplicity and potential for misclassifications of comorbidities in the EHR, these borderline associations need to be interpreted with caution.

**Fig 4.**
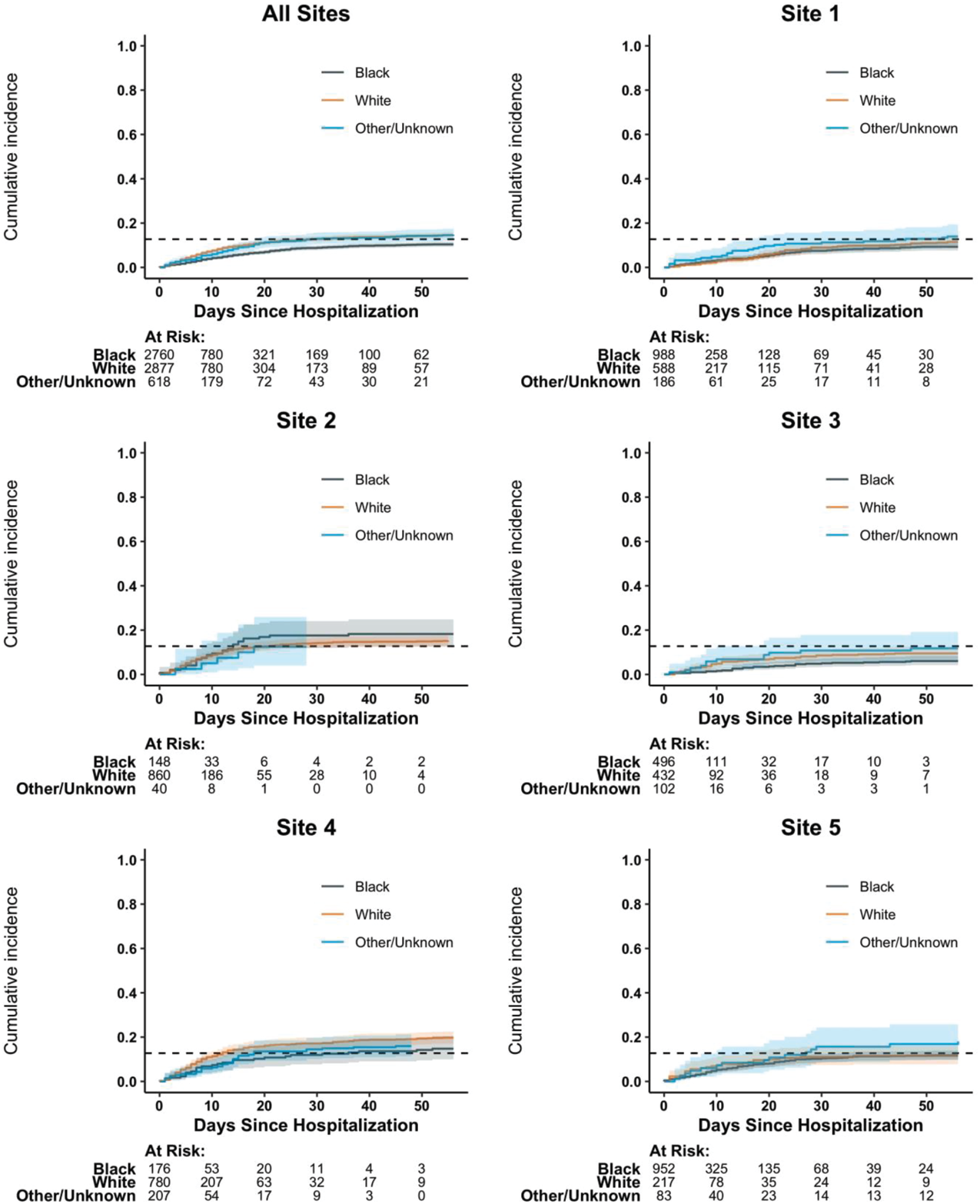
Cumulative incidence curves for death by admitting hospital and race. N=6255; Site 1 (n=1762), Site 2 (n=1048), Site 3 (n=1030), Site 4 (n=1163), Site 5 (n=1252) are unique hospitals in the University of Pennsylvania Health System.

## Discussion

Concurrent changes in patient demographics and health care provider practice patterns over the course of the pandemic complicate assessments of the role of individual risk factors on hospital outcomes. Clinical management of COVID-19 has continually evolved, in terms of what practices and therapies are employed and when patients receive ICU-level care. In our analysis of 6255 patients hospitalized with COVID-19 in the UPHS, we found that as time progressed, the percent of patients placed on mechanical ventilation decreased. Meanwhile, the mortality in ventilated patients increased while overall mortality decreased, suggesting this measure was increasingly reserved for a more severely ill patient population. In parallel, increased community spread over time led to admissions of younger and overall healthier individuals, which contributed to the drop in mortality rates mid-way through the pandemic; however, older age groups made up a larger percent of admissions during both winter periods. With potential new waves of infection, changes in circulating virus variants and expanded vaccine uptake will likely precipitate further shifts in the demographics and clinical outcomes of those acquiring infection and experiencing severe illness.

In addition to exploring trends over time, our analysis employed a novel multi-state model to investigate determinants of patient outcomes including death, discharge and hospital re-admission. This approach allowed us to capture the movement of patients between these states with more detail compared to the traditional approaches that have dominated the COVID-19 literature, which presented regression analyses for a single dichotomized (two-state) outcome such as 30-day mortality. Unlike the traditional two-state Cox or logistic model approach, the multi-state analysis also allows for data on multiple outcomes for a single patient, further allowing for nuanced assessments, such as directly modeling the number of prior admissions or risk factors affecting negative outcomes after discharge. The simplified, single outcome strategies have been used in similar COVID-19 cohort studies to show that age, severe illness at admission, existing comorbidities and socioeconomic factors may impact hospital outcomes [2,3,18]. Our analysis largely corroborated these results but also highlighted the nuanced associations with death, discharge, and re-hospitalization rates. There was appreciable variation in mortality rates by admitting hospital that could not be explained by the observed patient characteristics. These results are consistent with those of a recent study of nearly 40,000 patients from 955 US hospitals, which found that standardized 30-day mortality rates varied from 9.1 to 15.6% [8]; however, there were few identified factors associated with these differences.

Our analyses found that, at time of hospital admission, Black patients had more risk factors associated with poor outcomes compared to other races, including a higher proportion with EHR-documented comorbidities, residence in a low-income zip code, and requirement of ICU-level care at admission. Despite this finding, unadjusted comparisons of the mortality rates in Black patients compared to White patients found that Black race was associated with better outcomes. After adjusting for a wide set of demographic and clinical risk factors, however, we did not find an effect of race on death. There were no strong associations in multivariate models for the other outcomes (transitions). Notably, we found that ignoring admitting hospital in our analyses led to qualitatively different results regarding the impact of race. This is an example of Simpson’s Paradox [19], which occurred because there were imbalances in racial makeup across hospitals and the outcomes rates by hospital were very different. This type of confounding may at least partially explain why our finding that hospital mortality risk did not appreciably vary by race differed from other published reports, including a recent study of nearly 10,000 hospitalized patients in New York City that reported Black race was associated with a 30% reduction in hospital death compared to Whites [9]. Our results are consistent with a recent study by Asch et al. 2021 of 44,217 Medicare Advantage enrollees aged 18 or older who were admitted with COVID-19 to one of 1,188 hospitals across the US, which also found that unadjusted differences in mortality by race were significant but were undetectable after adjusting for admitting hospital [20]. In contrast to our study, however, Asch et al. [20] found that in their Medicare population, Black patients were admitted in larger proportion to hospitals with poorer outcomes. These investigators hypothesized that if Blacks were admitted to better performing hospitals, they would have better outcomes overall. Our results support this hypothesis. Further research is needed to understand whether public insurance status was also affecting the two racial groups differently.

Our results contribute to delineating the complex role of race and racial health disparities in patient exposures, pre-existing comorbidities, and outcomes from COVID-19. Several authors have discussed that structural factors are critical driving forces behind COVID-19 disparities and that system level reforms and interventions are required to fully address them [21, 22].

In contrast to race, Hispanic/Latinx ethnicity was associated with a decreased mortality hazard. This result should be interpreted with caution, however, given the relatively small number of Hispanic/Latinx individuals who were younger and had fewer comorbidities in our cohort. A key limitation of our analysis is the reliance on EHR, which could introduce confounding if the level of completeness was disproportionally affected by ethnicity.

Our findings corroborate the impact of COVID-19 in the elderly seen broadly. Patients over age 75 were at greatest risk for death and/or readmission. Improved strategies for post-discharge care of COVID-19 patients are needed to address the sustained vulnerability of these patients [23].

Passive data collection through routine electronic health records facilitates rapid research; however, the data are subject to a number of limitations. Records of comorbidities at hospital admission may be incomplete. Further, comorbidities diagnosed and treated outside the Penn health care system may not have been captured. Information regarding discharge destination (e.g. independent living or nursing home facility) was not available. Penn EHR may also have missed readmissions or deaths that occurred outside the Penn system. Linkage with federal and state registries will provide more complete data on survival, but these registries take time to be updated. Future work is needed to fully capture long-term outcomes of COVID-19.

## Data Availability

Data are not publicly available; however the primary author can be contacted to obtain information about the process necessary to obtain data.

## S1 Appendix Hospital descriptions and ICD-10 Code comorbidity definitions

### Section 1.1 Hospital descriptions^1^

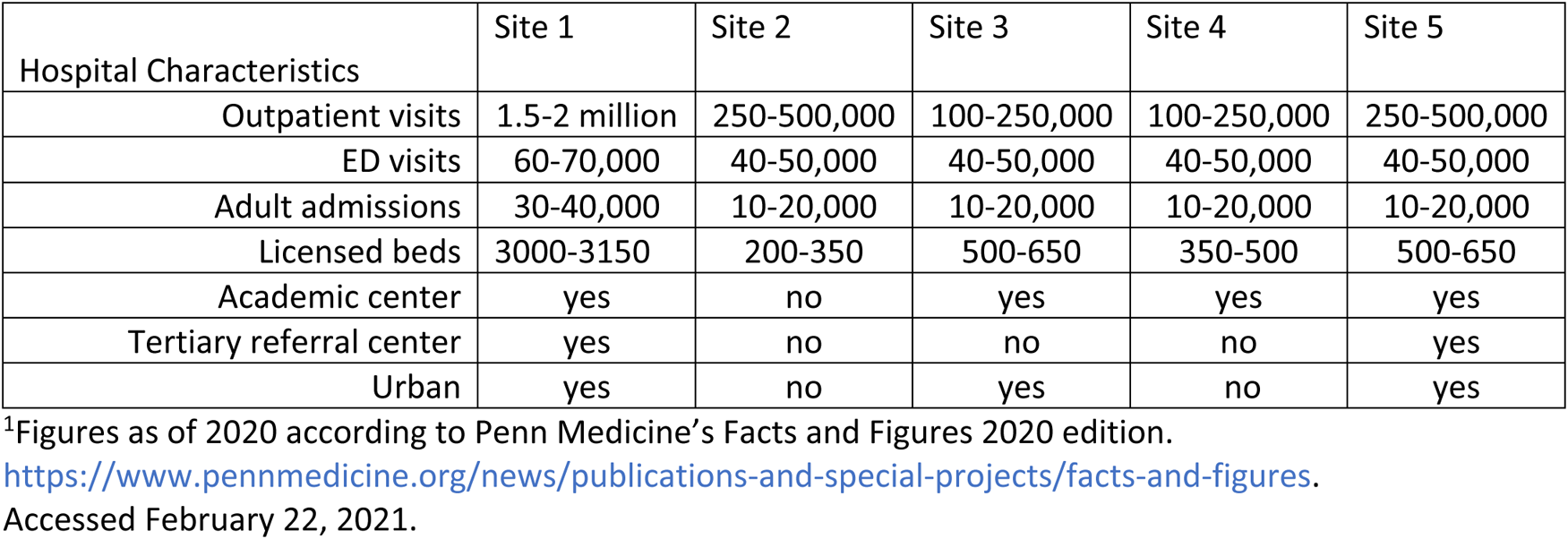

### Section 1.2 ICD-10 Code comorbidity definitions

| Condition | ICD10 Codes | Notes |
| --- | --- | --- |
| Diabetes | E08 – E13 | **“Diabetes mellitus.”** These are all the codes for diabetes mellitus excluding neonatal and gestational diabetes. |
| Diabetes Type I | E10 | **“Type 1 diabetes mellitus”** |
| Diabetes Type II | E11 | **“Type 2 diabetes mellitus”** |
| Obesity | E66 (excluding E66.3) | **“Overweight and Obesity.”** All sections of E66 code for obesity besides E66.3 which codes for overweight. We may also use given height and weight data or BMI (coded in Z68) |
| Chronic Kidney Disease | N18, N19 | **“Chronic kidney disease (CKD)”** and **“Unspecified kidney failure”** |
| Chronic Liver Disease | K70-K77 (excluding K71.2 and K72.0), B18- | **“Diseases of the liver.”** Also includes hemochromatosis (E83.11), Reye's syndrome (G93.7), viral hepatitis (B18-B19), Wilson's disease |
|  | B19, E83.11, G93.7, E83.0 | (E83.0). Excludes acute hepatitis (K71.2), acute and subacute hepatic failure (K72.0). |
| Chronic Respiratory Disease | J40-J99 (Excluding explicitly acute or COVID-induced) | **Includes multiple sections.** Includes all diseases of the respiratory system besides influenza and pneumonia (J09-J18), acute upper (J00-J06), acute lower (J20-J22) respiratory infections, and other diseases of upper respiratory tract (J30-J39). Includes chronic lower respiratory diseases (J40-47), lung diseases due to external agents (J60-J70), Other respiratory diseases principally affecting the interstitium (J80-J84), suppurative and necrotic conditions of the lower respiratory tract (J85-J86), Other diseases of the pleura (J90-J94), procedural complications and disorders of the respiratory system not elsewhere classified (J95), and other diseases of the respiratory system (J96-J99). Because COVID and related symptoms fall into many of these sections, we restricted diagnoses to those which were recorded at least one month prior to hospital admission for COVID. |
| Asthma | J45, J69.8, J82, J60, R06.2, J67.8 | **"Asthma."** All cases of asthma including detergent asthma (J69.8), eosinophilic asthma (J82), miner's asthma (J60), wheezing NOS (R06.2), wood asthma (J67.8). |
| Chronic Oxygen Requirement | Z99.81 | **"Dependence on supplemental oxygen."** Only included if recorded at least 30 days before hospital admission for COVID-19 to avoid categorizing COVID-19 oxygen treatment as a pre-existing condition. |
| Coronary Artery Disease | I25.1, I25.7, I25.81 | **"Coronary atherosclerosis."** These are the codes that the ICD10 says code for coronary atherosclerosis. |
| Hypertension | I10 | **"Essential (primary) hypertension."** Excludes primary hypertension of the brain (I60-I69) and eye (H35.0). Also excludes hypertensive heart (I11), chronic kidney (I12), heart and chronic kidney (I13), secondary hypertension (I15), and hypertensive crisis (I16). |
| Congestive Heart Failure | I50 | **"Heart failure."** The ICD10 doesn't specify congestive, but this seems to cover congestive heart failure. Includes all types of heart failure excluding cardiac arrest. |
| Cancer | C, D46 | **“Malignant neoplasms.”** Excludes benign neoplasms (D00-D36). Includes myelodysplastic syndromes (D46). |
| Immune Deficiency | B80-B84, B89, D20 | **“Immunodeficiency disorders.”** Includes immunodeficiency with predominantly antibody defects (D80), combined immunodeficiencies (D81), immunodeficiency associated with other major defects (D82), common variable immunodeficiency (D83), other immunodeficiencies (D84), HIV (B20), disorders involving the immune mechanism (D89). |
| Cardiovascular Disease | I00-I99 | **“Diseases of the circulatory system.”** Acute rheumatic fever (I00-I02), Chronic rheumatic heart diseases (I05-I09), hypertensive diseases (I10-I16), ischemic heart diseases (I20-I25), pulmonary heart disease and diseases of pulmonary circulation (I26-I28), other forms of heart disease (I30-I52), cerebrovascular diseases (I60-I69), diseases of arteries, arterioles, and capillaries (I70-I79), diseases of veins, lymphatic vessels and lymph nodes (I80-I89), other and unspecified disorders of the circulatory system (I95-I99). Excluded new (later than 30 days before hospital admission) diagnoses related to thrombosis and cardiac arrest in order to avoid misclassifying COVID-induced events as comorbidities. |

Data on pre-existing conditions in patients was gathered from ICD-10 codes documented in the past year based on the patient’s electronic health record in Penn Medicine. Conditions were grouped according to the conventions below. Respiratory diseases and chronic oxygen requirement were only included if the diagnosis occurred at least one month before the patient was admitted to the hospital with COVID-19 in order to avoid categorizing COVID symptoms as pre-existing conditions. **Bold** content is the title of the recommended sections as specified by the ICD10 tabular.

---

**S1 Fig.**
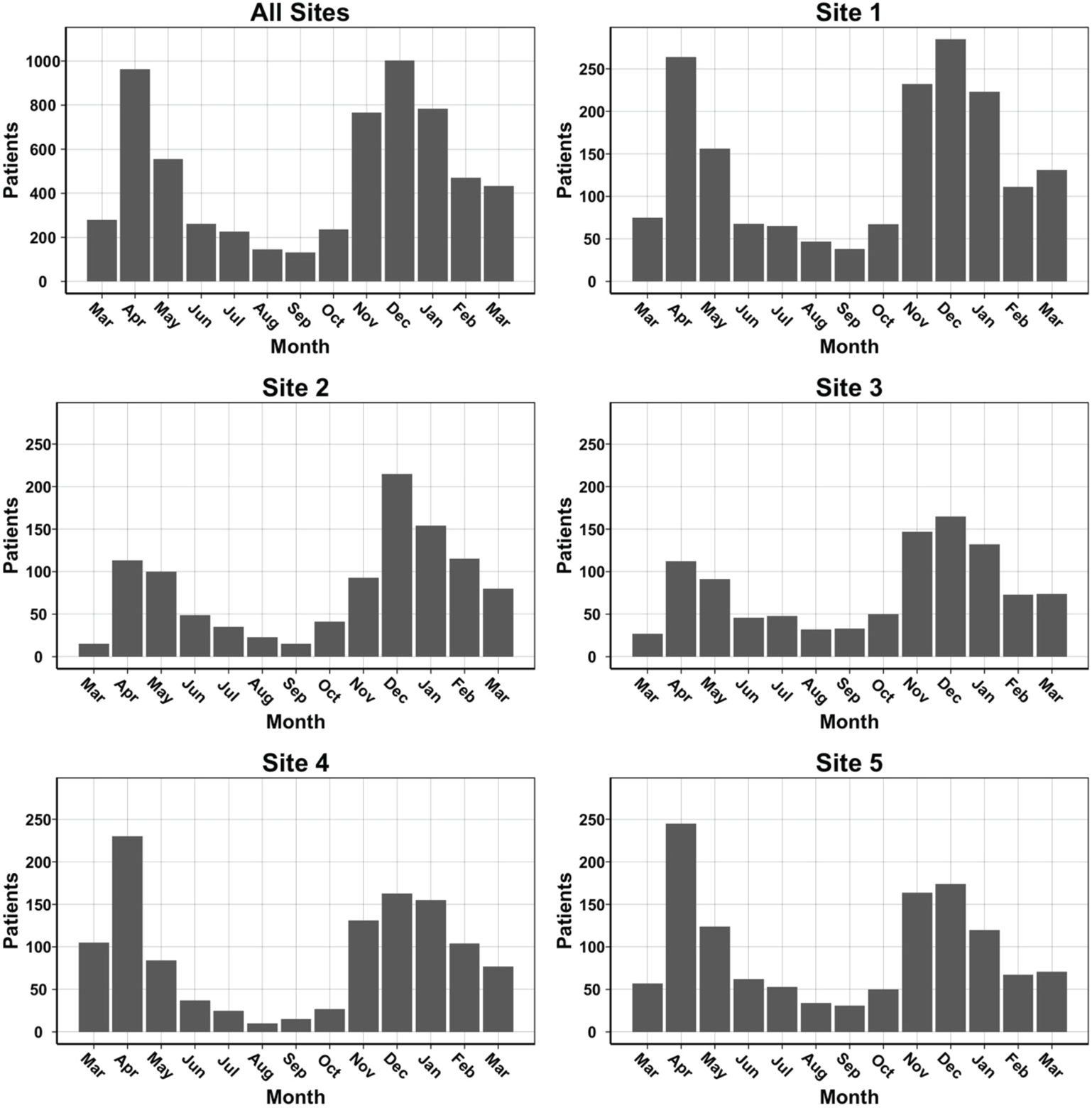
Number of unique patients admitted to a hospital in the UPHS with COVID-19 during each month from March 2020 to March 2021. Site 1 (n=1762), Site 2 (n=1048), Site 3 (n=1030), Site 4 (n=1163), Site 5 (n=1252) are unique hospitals in the University of Pennsylvania Health System.

**S2 Fig.**
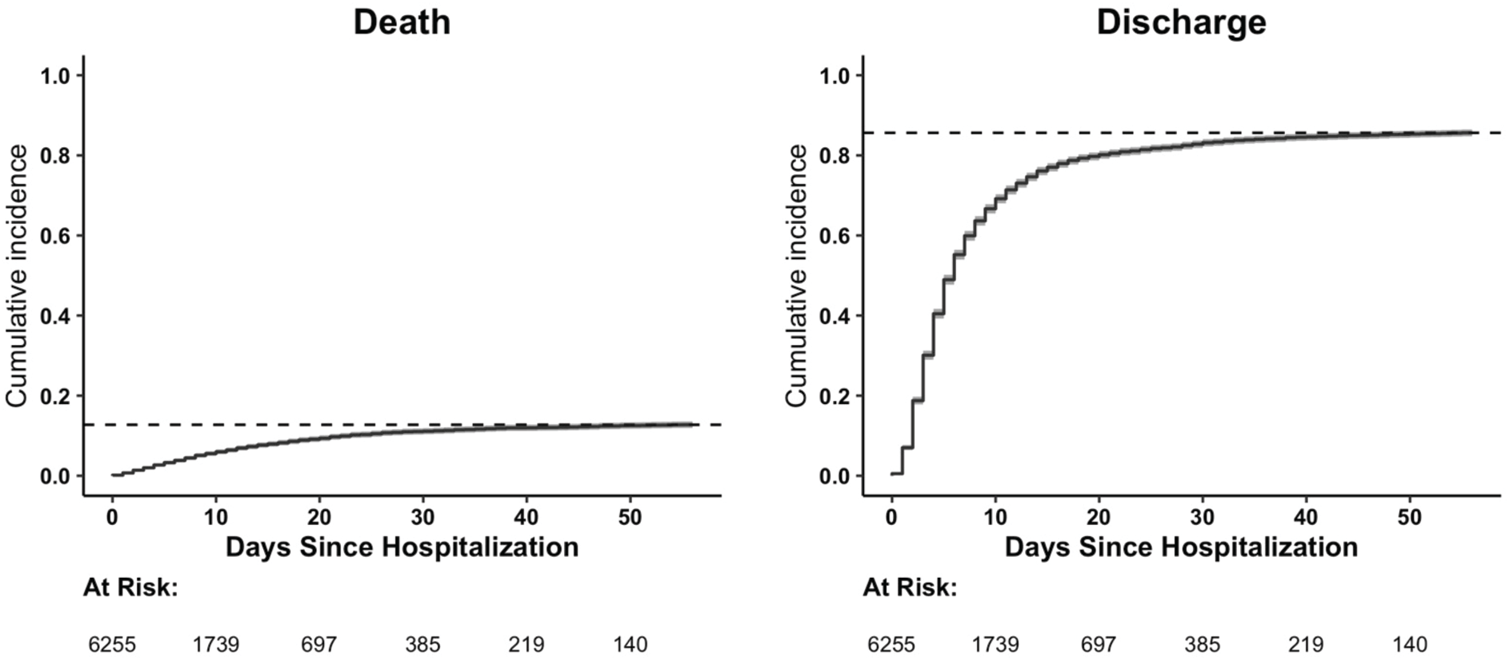
Cumulative incidence curves for death and discharge. N=6255.

**S3 Fig.**
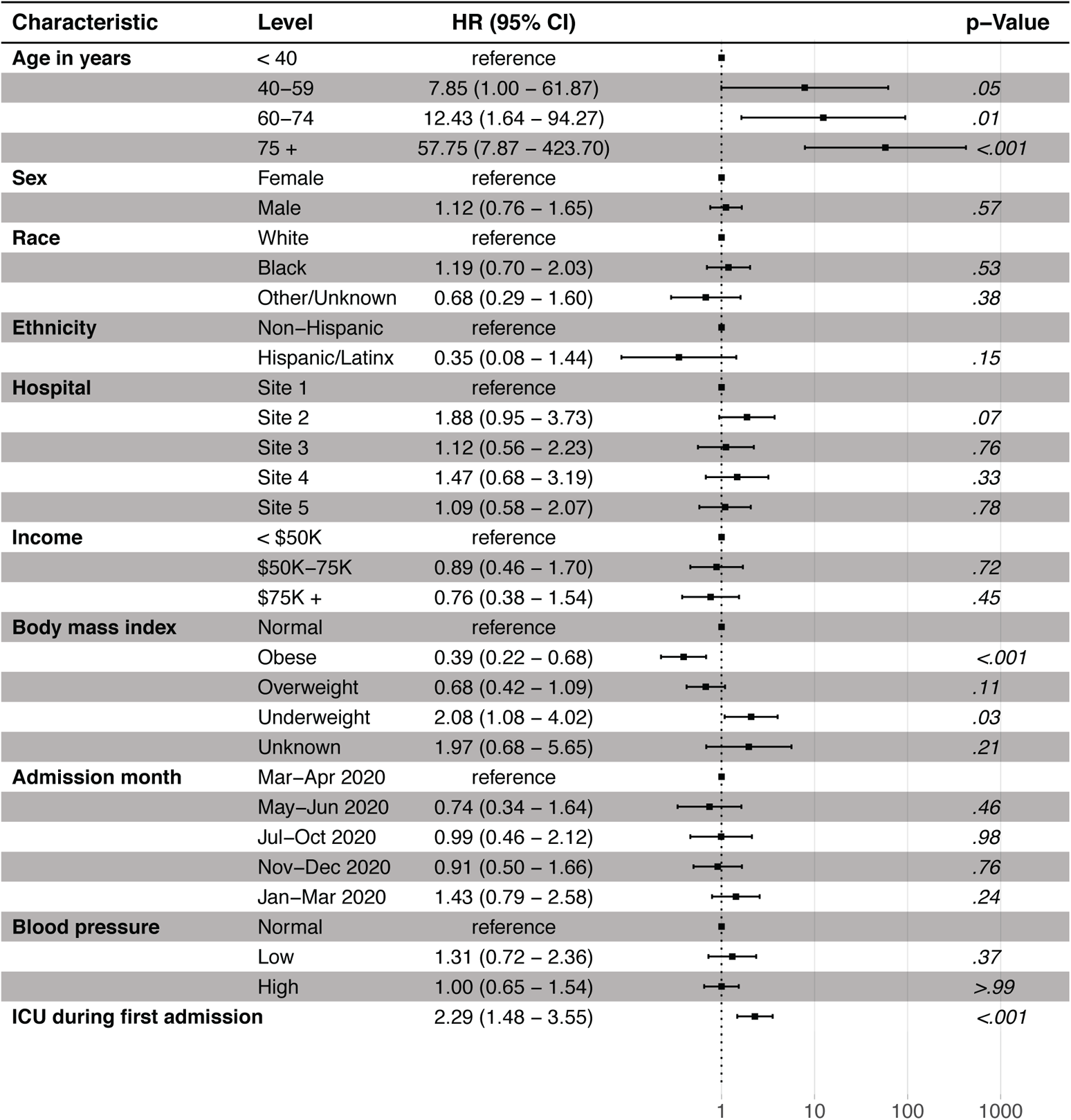
Hazard ratio and 95% CI for cause-specific Cox regression model for discharge to death transition adjusted for disease severity and other patient characteristics at admission (model 1). N=6233. 22 observations deleted due to missingness in zip code (15) and blood pressure (7). Income: Median Household Income in patient’s 5-digit zip code, as determined by the 2014-2018 5-year ACS; CI: Confidence interval; Site 1 (n=1762), Site 2 (n=1048), Site 3 (n=1030), Site 4 (n=1163), Site 5 (n=1252) are unique hospitals in the University of Pennsylvania Health System.

**S4 Fig.**
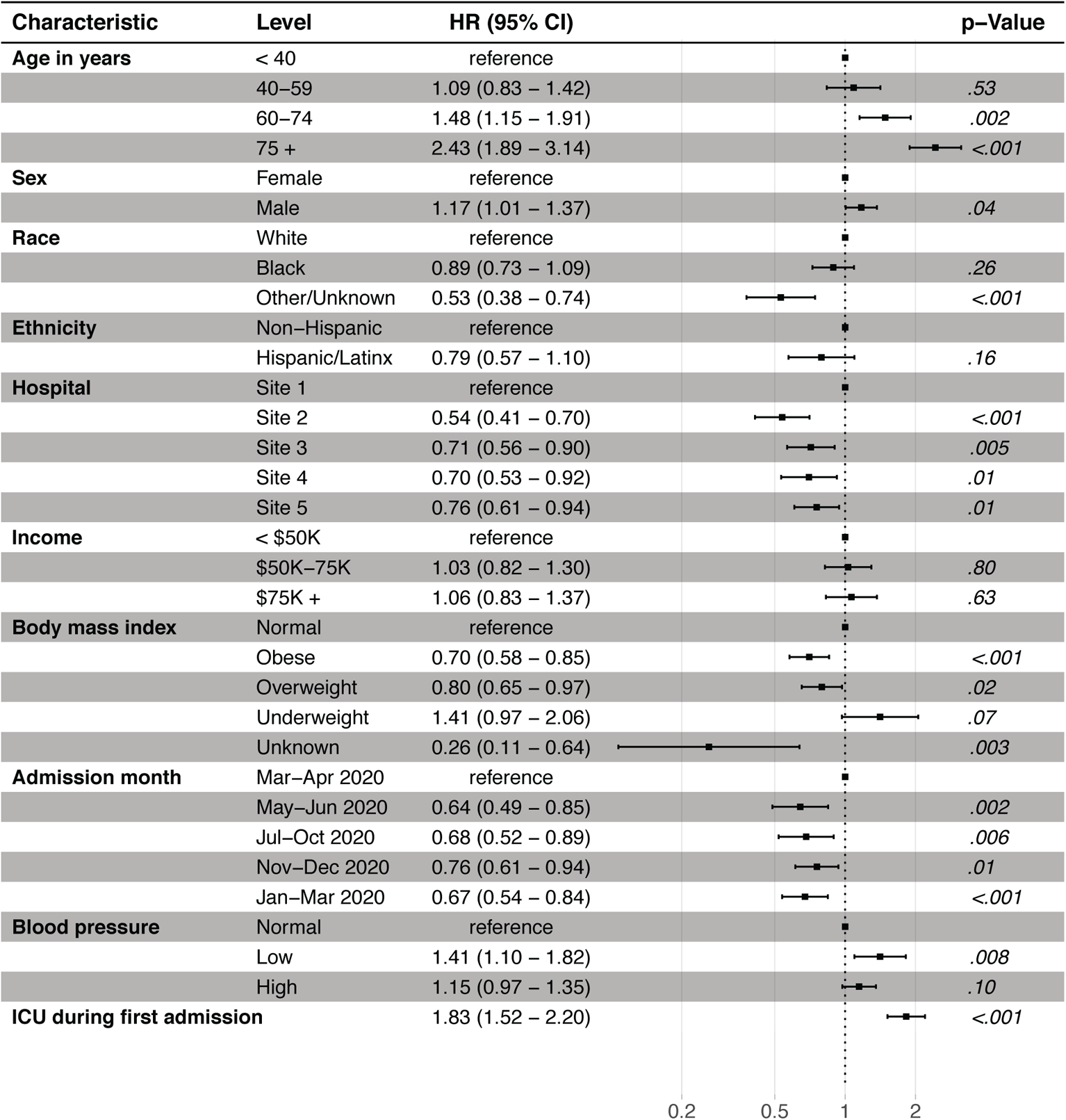
Hazard ratio and 95% CI for cause-specific Cox regression model for discharge to re-hospitalization transition adjusted for disease severity and other patient characteristics at admission (model 1). N=6233. 22 observations deleted due to missingness in zip code (15) and blood pressure (7). Income: Median Household Income in patient’s 5-digit zip code, as determined by the 2014-2018 5-year ACS; CI: Confidence interval; Site 1 (n=1762), Site 2 (n=1048), Site 3 (n=1030), Site 4 (n=1163), Site 5 (n=1252) are unique hospitals in the University of Pennsylvania Health System.

**S5 Fig.**
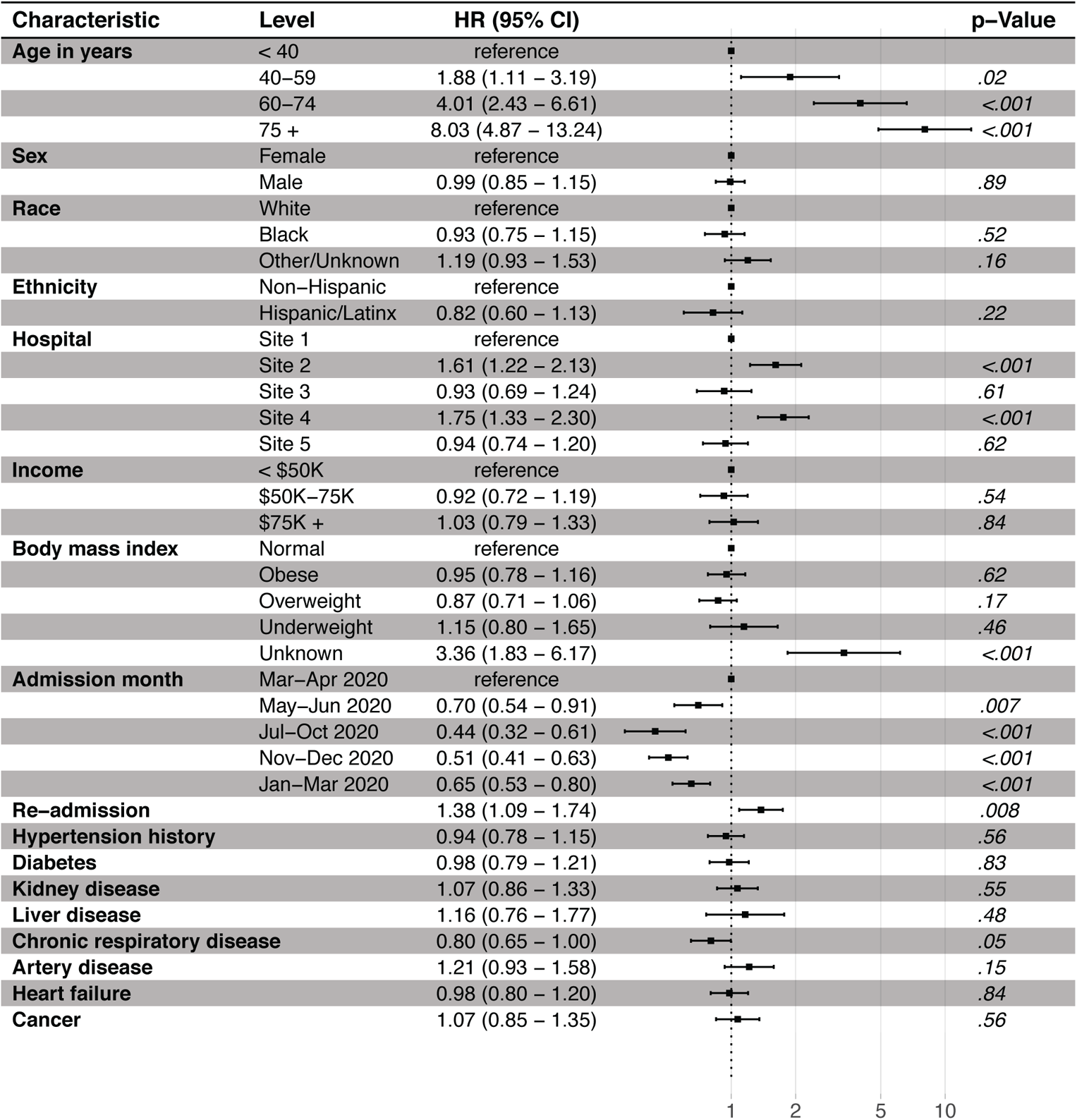
Hazard ratio and 95% CI for cause-specific Cox regression model for hospital to death transition adjusted for comorbidities and other patient characteristics at admission (model 2). N=6240. 15 observations deleted due to missingness in median household income. ICU Stay: Indicator for whether a patient spent any time in the ICU during first hospital stay; CI: Confidence interval; Site 1 (n=1762), Site 2 (n=1048), Site 3 (n=1030), Site 4 (n=1163), Site 5 (n=1252) are unique hospitals in the University of Pennsylvania Health System.

**S6 Fig.**
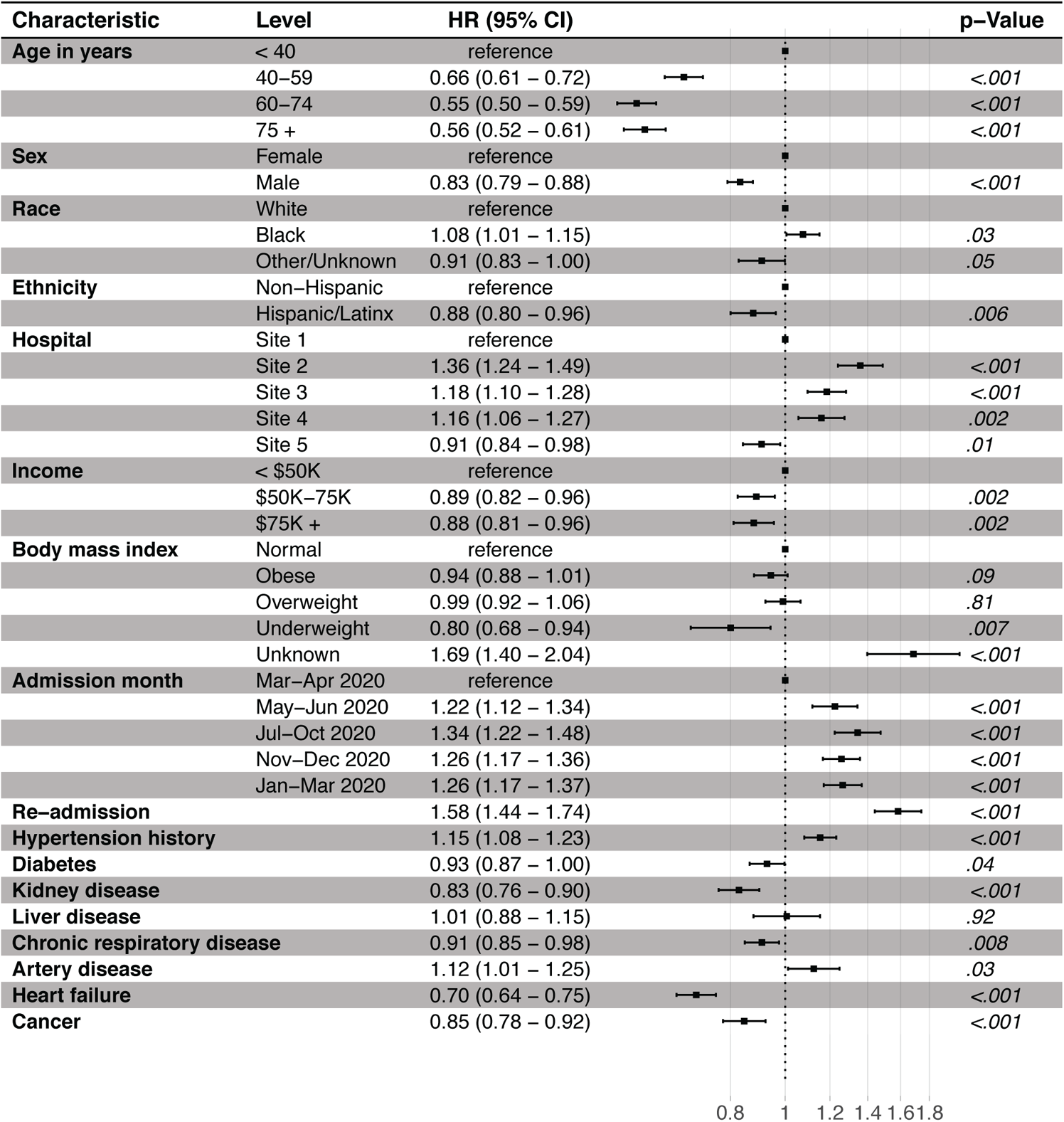
Hazard ratio and 95% CI for cause-specific Cox regression model for hospital to discharge transition adjusted for comorbidities and other patient characteristics at admission (model 2). N=6240; 15 observations deleted due to missingness in median household income. BMI: Body mass index; CI: Confidence interval; Site 1 (n=1762), Site 2 (n=1048), Site 3 (n=1030), Site 4 (n=1163), Site 5 (n=1252) are unique hospitals in the University of Pennsylvania Health System.

**S7 Fig.**
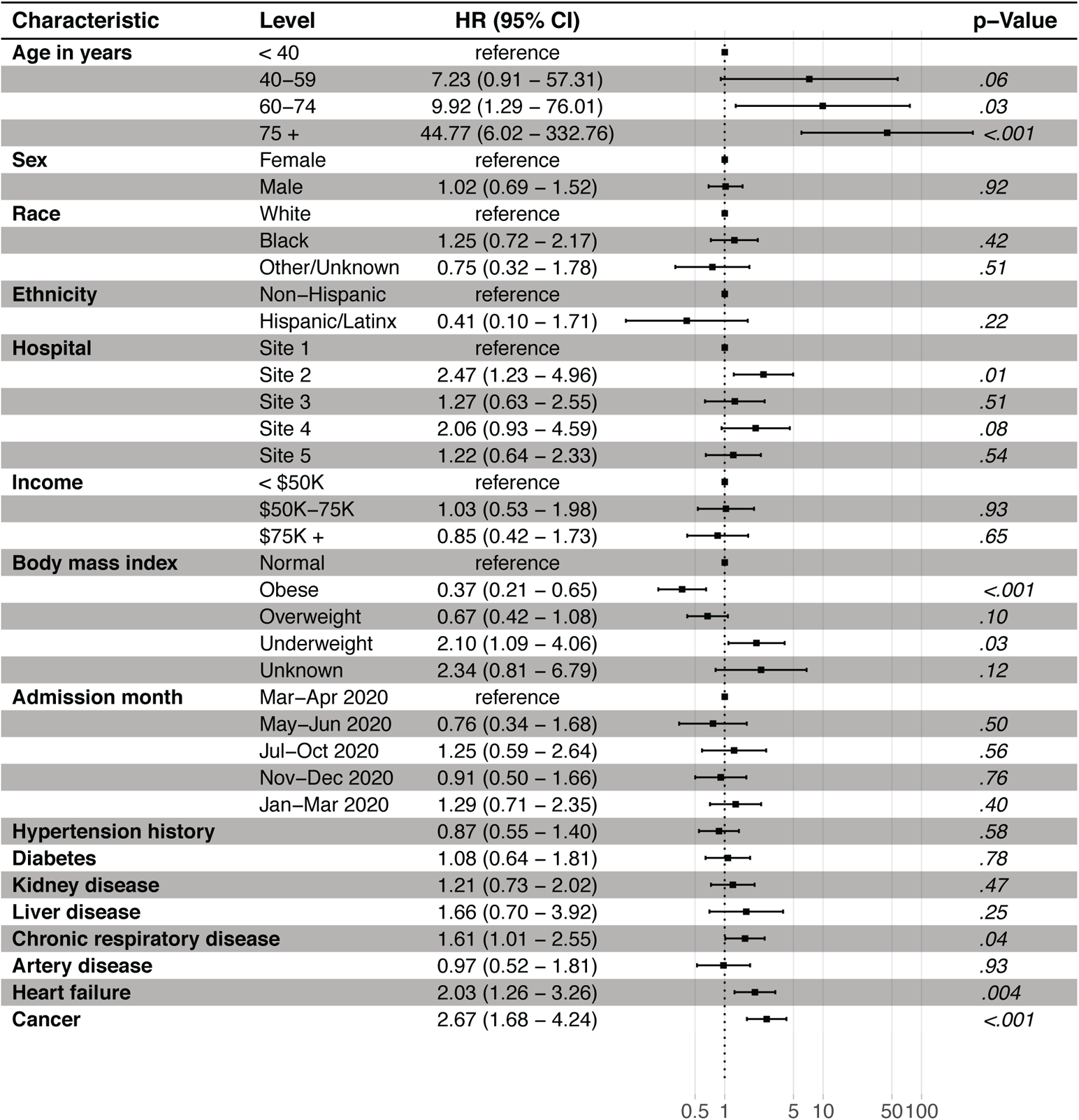
Hazard ratio and 95% CI for cause-specific Cox regression model for discharge to death transition adjusted for comorbidities and other patient characteristics at admission (model 2). N=6240; 15 observations deleted due to missingness in median household income. BMI: Body mass index; CI: Confidence interval; Site 1 (n=1762), Site 2 (n=1048), Site 3 (n=1030), Site 4 (n=1163), Site 5 (n=1252) are unique hospitals in the University of Pennsylvania Health System.

**S8 Fig.**
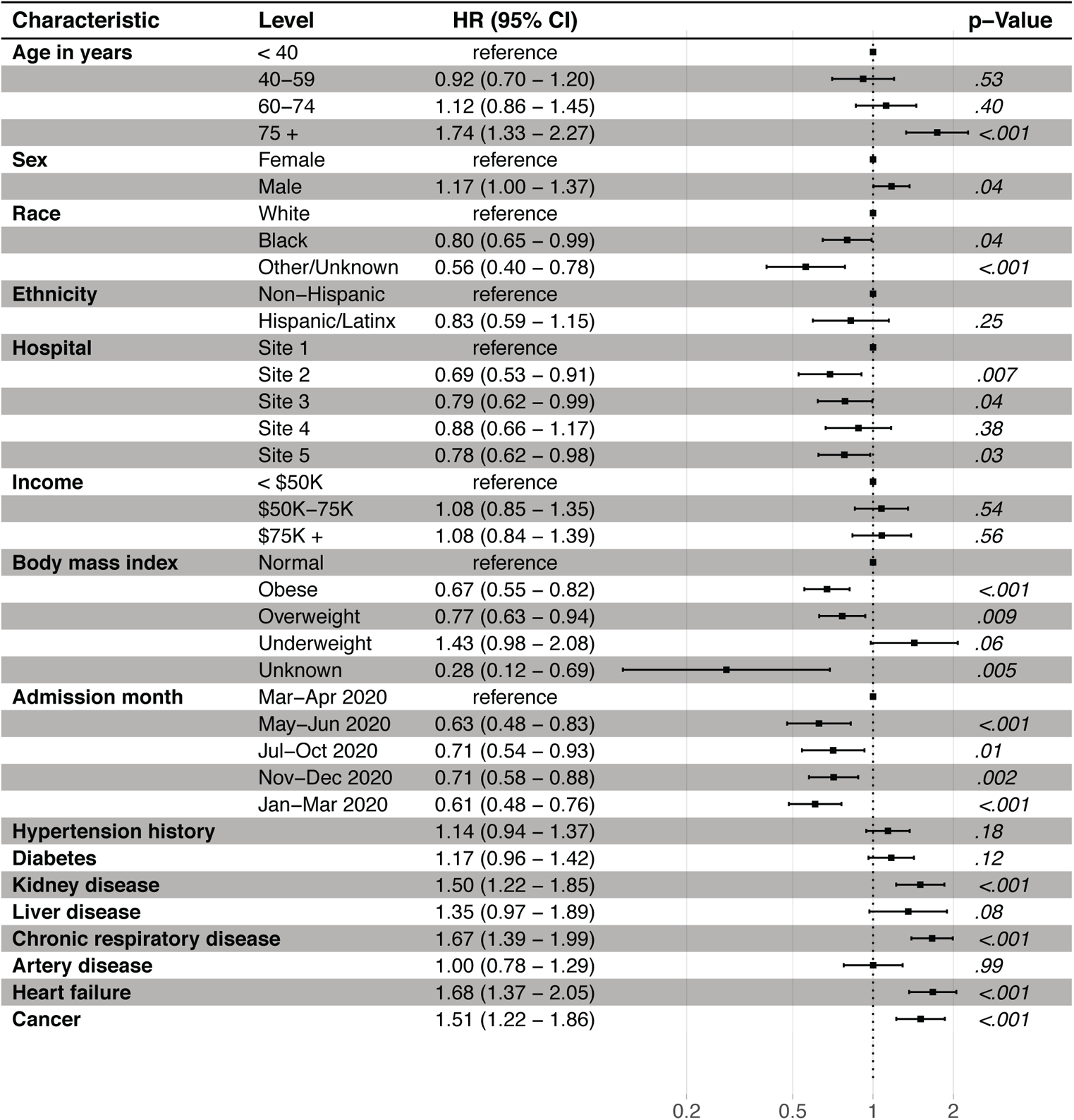
Hazard ratio and 95% CI for cause-specific Cox regression model for discharge to re-hospitalization transition adjusted for comorbidities and other patient characteristics at admission (model 2). N = 6240; 15 observations deleted due to missingness in median household income. BMI: Body mass index; CI: Confidence interval; Site 1 (n=1762), Site 2 (n=1048), Site 3 (n=1030), Site 4 (n=1163), Site 5 (n=1252) are unique hospitals in the University of Pennsylvania Health System.

**S9 Fig.**
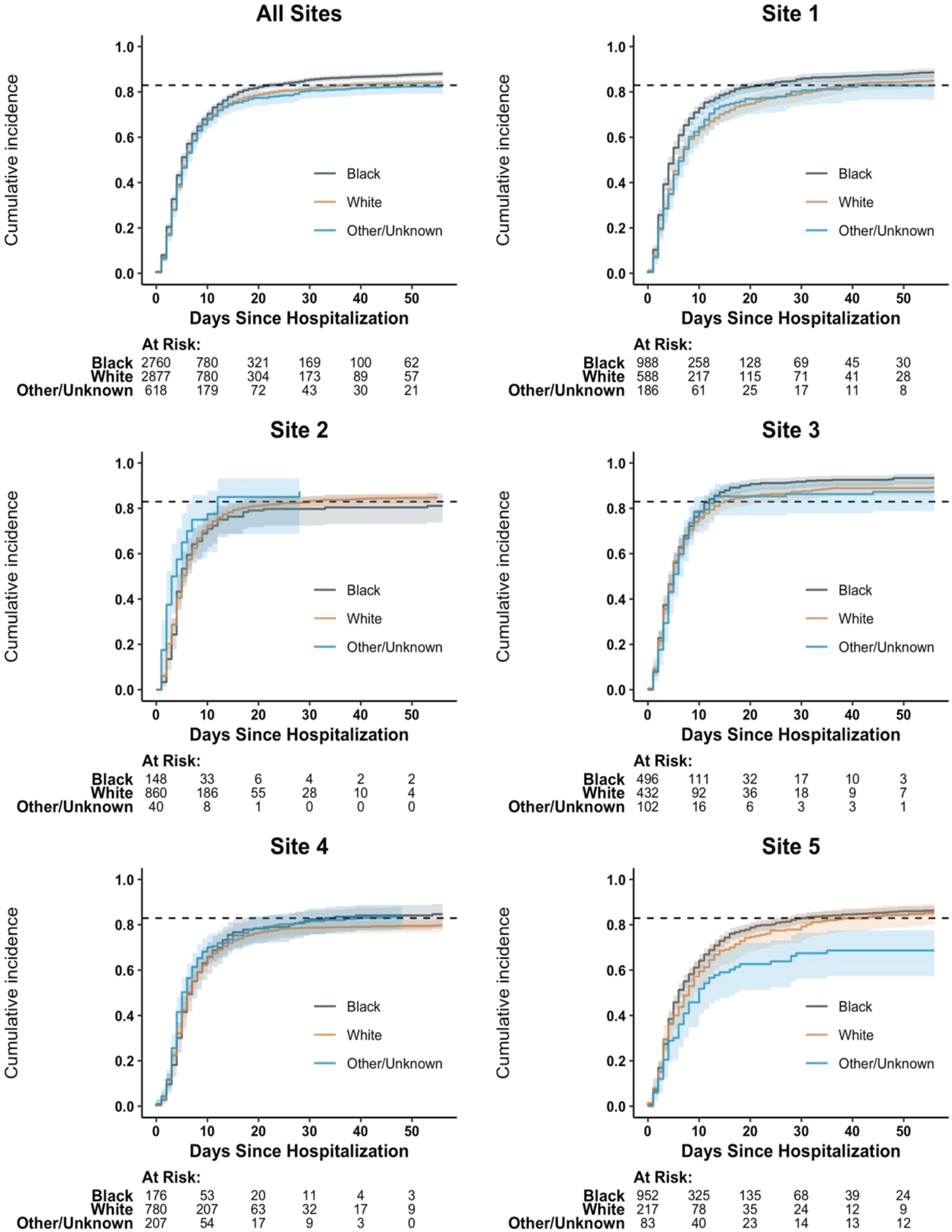
Cumulative incidence for discharge by admitting hospital and race. N=6255. Site 1 (n=1762), Site 2 (n=1048), Site 3 (n=1030), Site 4 (n=1163), Site 5 (n=1252) are unique hospitals in the University of Pennsylvania Health System.

**S1 Table.** Comorbidity at COVID-19 hospital admission by race. N=6255. Percentages may not add up to 100% due to missing data in median household income or rounding elsewhere. P-values from chi-squared test. Any ICD-10 comorbidity included diabetes, cardiovascular disease, respiratory disease, kidney disease, liver disease, immune deficiency, and chronic oxygen requirement. See Supplemental S1 Appendix for more details.

| Condition |  | White<br>(N = 2877) | Black<br>(N = 2760) | Other<br>(N = 618) | P-Value |
| --- | --- | --- | --- | --- | --- |
| Any ICD-10 comorbidity |  | 1627 (56.6) | 1967 (71.3) | 298 (48.2) | < 0.001 |
| Diabetes |  | 386 (13.4) | 789 (28.6) | 65 (10.5) | < 0.001 |
| Kidney disease |  | 223 (7.8) | 515 (18.7) | 41 (6.6) | < 0.001 |
| Liver disease |  | 94 (3.3) | 104 (3.8) | 13 (2.1) | 0.107 |
| Respiratory disease |  | 398 (13.8) | 556 (20.1) | 59 (9.5) | < 0.001 |
| Cardiovascular disease |  | 1353 (47.0) | 1659 (60.1) | 240 (38.8) | < 0.001 |
| Heart failure |  | 306 (10.6) | 443 (16.1) | 65 (10.5) | < 0.001 |
| Cancer |  | 319 (11.1) | 273 (9.9) | 32 (5.2) | < 0.001 |
| ICU at admission |  | 391 (13.6) | 420 (15.2) | 116 (18.8) | 0.003 |
| Age in years | < 40 | 415 (14.4) | 589 (21.3) | 113 (18.3) | < 0.001 |
|  | 40-59 | 663 (23.0) | 811 (29.4) | 170 (27.5) |  |
|  | 60-74 | 893 (31.0) | 830 (30.1) | 198 (22.2) |  |
|  | 75 + | 906 (31.5) | 530 (19.2) | 137 (22.2) |  |
| Income | <\$50K | 393 (13.7) | 1997 (72.4) | 170 (27.5) | < 0.001 |
| | \$50K-\$75K | 682 (23.7) | 428 (15.5) | 141 (22.8) | |
| | \$75K+ | 1795 (62.4) | 329 (11.9) | 305 (49.4) | |
| Body mass index (BMI) | Normal | 675 (23.5) | 526 (19.1) | 194 (31.4) | < 0.001 |
|  | Obese | 1166 (40.5) | 1410 (51.1) | 191 (30.9) | < 0.001 |
|  | Overweight | 888 (30.9) | 691 (25.0) | 205 (33.2) |  |
|  | Underweight | 93 (3.2) | 72 (2.6) | 11 (1.8) |  |
|  | Unknown | 55 (1.9) | 61 (2.2) | 17 (2.8) |  |

**S2 Table.**
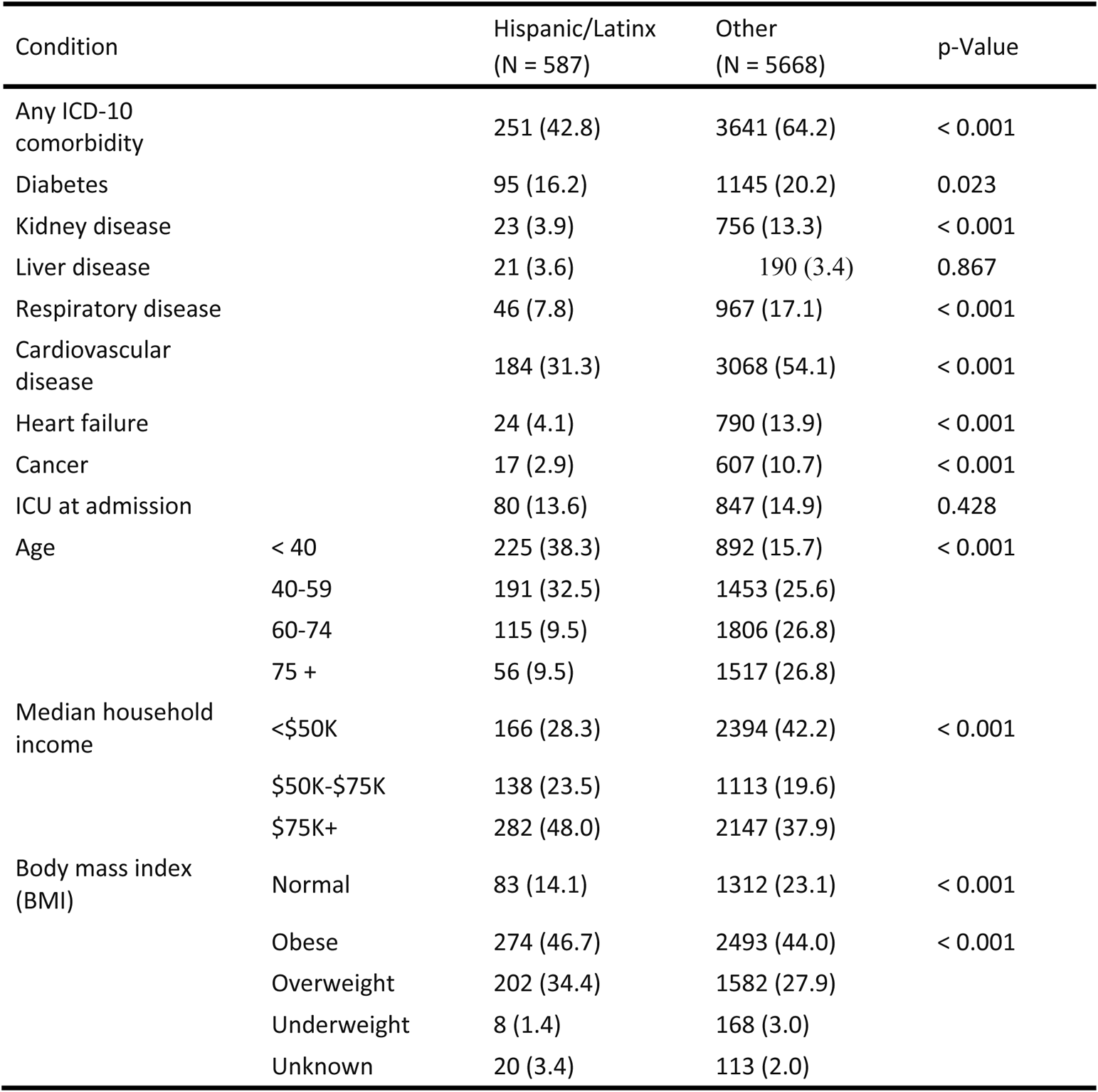
Comorbidity at COVID-19 hospital admission by Hispanic/Latinx ethnicity. N=6255. Percentages may not add up to 100% due to missing data in median household income or rounding elsewhere. P-values were for the chi-squared test. Any ICD-10 comorbidity included diabetes, cardiovascular disease, respiratory disease, kidney disease, liver disease, immune deficiency, and chronic oxygen requirement. See Supplemental S1 Appendix for more details.

